# Genotoxicity evaluation of levonorgestrel-releasing intrauterine system (LNG-IUS) in exfoliated cervical cells using the micronucleus (MN) test

**DOI:** 10.1101/2021.07.08.21260031

**Authors:** Ayse Gul Zamani, Rengin Karatayli, Gezginc Kazim, Aynur Acar, Mahmut Selman Yildirim

## Abstract

**Purpose:** This study aimed to determine whether any relationships exist between the levonorgestrel-releasing intrauterine system (LNG-IUS) and micronuclei or other nuclear anomalies, including condensed chromatin, karyorrhexis, and karyolysis, on the cervical epithelium in young women.

**Methods:** A prospective observational study was conducted. The study population comprised healthy women aged ≤40 years who were referred for birth control. Cervical smears that were obtained from 18 women before and three months after LNG-IUS insertion were tested for micronuclei and other nuclear anomaliesusing the micronucleus test.

**Results:** The results revealed no statistically significant difference (P>0.05) in the frequency of micronucleated exfoliated cervical mucosa cells after LNG-IUS exposure. However, LNG-IUS was able to increase other nuclear alterations closely related to cytotoxicity.

**Conclusions:** Data indicated that exposure to LNG-IUS may not be a factor in inducing chromosomal damage, but it can promote cytotoxicity.

## Introduction

The levonorgestrel-releasing intrauterine system (LNG-IUS) known as Mirena was first registered in Finland in 1990. It is an effective and widely used form of contraception. The LNG-IUS releases approximately 20 μg levonorgestrel daily into the uterine cavity. In addition to contraceptive purposes, it is now commonly used in the management of heavy menstrual blood loss.The LNG-IUS causes endometrial glands to become inactive, leading to stromal decidualization, and these morphological changes continue until the LNG-IUS is removed (Silverberg et al. 1986). Some studies have not found a relationship between the use of intrauterine devices (IUD) and cervical cancer (Sandmire et al. 1976; Lassise et al. 1991; Li et al. 2000; Roura et al. 2016; Curtis et al. 2007). However, other studies have showed that there may be a possible link between hormonal contraceptive use and cervical cancer risk, particularly with long durations of combined hormonal (≥10 years) or injectable contraceptive use (≥5 years).

These studies are limited, and the link between IUD usage and the development of precancerous cervical lesions and cervical cancer remains poorly understood (Smith et al. 2003; Appleby et al. 2007). Themicronucleusassaydetectsmicronuclei, whichserve as a highlysensitive marker of DNA damage. MN are whole chromosomes or acentric fragments that are monitored outside the nucleus. This test is a minimally invasive method for monitoring genetic damage and investigating the genotoxicity of different chemical substances (Thomas and Fenech 2011).The MN assay evaluates the magnitude of DNA damage and chromosomal instability produced by low doses of carcinogenic substances (Thomas and Fenech 2011; Suhas et al. 2004). Studies have shown that MN is a useful marker for detecting a predisposition for cancer (Chatterjee et al. 2009; Bolognesi et al. 2015). The direct effect of LNG-IUS on cervical cells and cancer has not been well studied. Therefore, this study evaluated the association between LNG-IUS use and cervical genotoxicity and cytotoxicity using the MN test.

## Materials and methods

### Selection and description of participants

A prospective observational study was conducted. The study population comprised 18 healthy women aged ≤40 years who were referred for birth control. Cervical smears were obtained from the participants before and three months after 52-mg LNG-IUS insertion and were tested for micronucleated cells and other nuclear alterations closely related to genotoxicity using the MN test.

All participants answered a modified version of the questionnaire of the Commission for Protection against Environmental Mutagens and Carcinogens (Carrano and Natarajan 1988). Information about contraceptive methods used, histories of sexually transmitted diseases, use of hormone replacement therapy, and the patients’ habits (smoking, drug use, and numbers of sexual partners) were obtained.

The study was approved by the Ethics Committee of Necmettin Erbakan University Meram Medical Faculty, Konya (protocol number: 2012/130). After the patients received and signed informed consent, exfoliated cervical cells were collected.

Exfoliated cervical cells were collected using Ayre spatulas and transferred to falcon tubes containing 0.9% physiologic serum for MN tests. After centrifugation, the supernatant was discarded, leaving the exfoliated cells in the pellet. Cells were treated with hypotonic solution for 5 min. Then, they were treated twice with 5 mL of a fresh mixture of cold methanol: aceticacid (3:1). Drops of the samples were placed on cold damp slides and allowed to dry. Samples were stained using 5% Giemsa for 5 min.

The slides were analyzed, and 2000 epithelial cells were counted at a magnification of 1000× (objective = 100×; with eyepiece = 10×). MN were determined according to the criteria by Stich and Rosin (Stich and Rosin 1983). Within the samples, only non-overlapping individual cells without folds were analyzed. MN were counted if the structures had a regular border, were located inside the cytoplasm, showed a staining intensity of less than or equal to that of the main nucleus, and were smaller than two-thirds of the size of the main nucleus. For cytotoxicity, the nuclear alterations of pyknosis, karyolysis, and karyorrhexis were considered according to the criteria used by Tolbert et al. (Tolbert et al.1992). The frequency of cells with MNand other nuclear anomalies, which are considered as markers of cytotoxic effects (condensed chromatin, karyorrhexis, pyknosis, and karyolysis), are reported in the results.

### Statistical methods

The Wilcoxon and Mann Whitney-U tests for dependent samples were used to compare the frequencies of MN and other cellular alterations related to cellular death among the samples before and after LNG-IUS exposure by using SPSS (Statistical Package for Social Sciences) software (version 22; SPSS, Chicago, IL, USA). Variables are displayed as the mean ± standard deviation in the tables. The significance level of all tests as considered as < 0.05.

## Results

The baseline characteristics of women who underwent LNG-IUS insertion are shown in Table 1. Before LNG-IUS insertion, the mean frequency of MNs was determined to be 0.04%. No statistically significant differences (P> 0.05) were observed after three months using the therapy. However, increases in other nuclear alterations (condensed chromatin, karyorrhexis, pyknosis, and karyolysis) were observed after LNG-IUS insertion. These data are summarized in Table 2.

**Table 1.** Baseline characteristics of women with LNG-IUS insertion.

| | Mean ± SD |
| --- | --- |
| Mean age (years) | 26± 4.06 (20-32) |
| Mean gravidity | 2.2±0.67 (1-3) |
| Mean parity | 2±0.65 (1-3) |
| Duration of sexual life<sup>a</sup> | 4.8±3.6 years |
| Body mass index | 24.75 ± 3,29 |

<sup>a</sup>Duration of sexual life in years from initial coitus. SD, standard deviation.

**Table 2.** Frequency of MN and other nuclear anomalies in exfoliated cervical cells of healthy females before and three months after LNG-IUS insertion.

| Groups | MN(%) |  | Other nuclear alterations*(%) |  |
| --- | --- | --- | --- | --- |
|  | No.of individuals | Mean±SD | No.of individuals | Mean±SD |
| Prior to LNG-IUS insertion | 18 | 0.04±0.04 | 18 | 5.82±4.14 |
| After LNG-IUS insertion | 18 | 0.05±0.07 | 18 | 12.46±3.24 |
|  |  | P = 0.6021 |  | P < 0.0001 |

\*Karyorrhexis, pyknosis, condensed chromatin and karyolysis; MN, micronuclei; SD, standard deviation.

## Discussion

The 52-mg LNG-IUS (Mirena, Bayer Healthcare, Whippany, NJ, USA), which carries levonorgestrel and releases the hormone slowly into the intrauterine cavity, is accepted as a gold-standard contraceptive IUDs are a reversible, long-term contraceptive method used by more than 100 million women worldwide. Mirena is also used for its protective effect in estrogen replacement therapy and for menorrhagia treatment (Grandi et al. 2018). Upon insertion, Mirena begins to gradually release small amounts of levonorgestrel. It is effective against implantation, sperm viability and movement, ovulation, and cervical secretion. Additionally, Mirena thickens the mucus in the cervical canal (Grandi et al. 2018; Nelson 2017). The existence of a possible relationship between LNG-IUS use and cervical cancer will affect women’s health globally. Twolarge meta-analysis studies revealed a low risk of cervical cancer associated with IUD use (Castellsagué et al. 2011, Cortessis et al. 2017),while other studies found no relationship between IUD use and cervical cancer (Curtis et al. 2007; Lassise et al. 1991; Roura et al. 2016; Sandmire et al. 1976; Smith et al. 2003). These studies were limited because IUD exposure has never been classified. Only one of these studies reported on the various types of IUDs, including inert and copper-containing devices. Since few studies have been conducted and IUDs are not classified, little is known about the relationship between precancerous cervical lesions and the development of cervical cancer and IUDs.

Particularly, it is not yet understood whether levonorgestrel-releasing IUDs play a role in the development of cervical cancer. Averbach et al. showed a relationship between IUD use, cervical pre-cancer (cervical intraepithelial neoplasia [CIN] 2, CIN3, adenocarcinoma in situ/CIN2+), and cancer (CIN3+). When theauthors classified their data by IUD type, they found that LNG-IUS use was related to CIN2+, but not to CIN3+ (Averbach et al. 2018). Hence, LNG-IUS use was associated with CIN2, which is considered a temporary infection, but not CIN3, which is defined as a high-grade pre-cancer. This finding suggests that there was no clinically significant harmful effect (Averbach et al. 2018). In a retrospective population-based cohort study that compared the relationship between contraceptive use and CIN3+ development, it was shown that oral contraceptive users had a higher risk of developing CIN3+ and cervical cancer than did IUD users. In another study, Spotnitz et al. compared copper IUD users and LNG-IUS users and revealed that the former group had a lower risk of high-grade cervical neoplasms (Spotnitz et al. 2020).

The MNcytome assay is a valuable test that evaluates genotoxicity and cytotoxicity at the chromosomal level using biomarkers. In the MN test, micronucleated cell and cells with other nuclear anomalies are evaluated together. The presence of mikronuclei, binucleated cells, and nuclear buds(“broken egg”)mostly indicate genotoxic events, while the frequency of nuclear anomalies such as pyknosis, karyolysis, condensed chromatin, and karyorrhexis are considered as cytotoxic effects (Tolbert et al.1992; Tak et al. 2014; Flores-Garcia et al. 2014). Cytotoxicity is defined as the potential of a compound to cause cell death, and subjecting human cells to cytotoxic compounds can lead to various cell fates, such as necrosis or apoptosis. However, not every cytotoxic agent has a genotoxic effect (Tak et al. 2014; Flores-Garcia et al. 2014). According to our results, although there was no change in MN frequencies, a statistical difference was found in the frequency of other nuclear abnormalities. Considering the results of the abovementioned studies as well as our results, LNG-IUS should not be considered to have a genotoxic effect. However, Hughes et al. reported cytological findings in the cervical smear of a 54-year-old Caucasian woman that was evaluated two years afterLNG-IUS placement (Hughes et al. 2005). The authorsstated that the nuclear chromatin was finely granular with occasional nuclear irregularity, and the nuclei contained one or more prominent nucleoli. They also described nuclear anomalies in a different manner such as condensed chromatin. In another study, Pap smears performed before and one year after LNG-IUS use revealed an increase in vaginal infections after one year (Donders et al. 2011). The human papillomavirus (HPV) is considered the main etiological agent of cervical cancer (zurHausen 2000), andLNG-IUS exposure may predispose the cervix to HPV infection. Invitro studies have shown that progesterone increases E6/E7 oncogene transcription in HPV-16-integrated celllines (Newfield et al.1998; Yuan et al. 1999). E6/E7 oncoproteins stimulate P53 degradation and increase the ability of viral DNA to transform cells and induce carcinogenesis (Yim et al. 2005). Thus, progesterone may be an additional preparatory factor in HPV-related malignancies. Furthermore, increased matrix metalloproteinase activity and angiogenesis may accelerate dysplasia in women using LNG-IUS approximately six months after placement. For these reasons, presumably, the authors wanted to draw attention to the smear findings. Some studies highlight the relationship between LNG-IUS and cancer (Loopik et al. 2020; Roura et al. 2016; Smith et al. 2003). Authors may also focus on this potential relationship due to the aforementioned concerns. We did not observe a genotoxic effect by LNG-IUS in the MN analysis performed before and three months after placement, but we suggest that there may be a cytotoxic effect. The short time between the first and last evaluationsisa limitation of our study. In a long-term follow-up study, the cytotoxic effect may becomea carcinogenic effect.

In conclusion, our results showed that the LNG-IUS Mirena is not a genotoxic agent, but it has a cytotoxic effect. HPV infections and the increased progesterone activity in a local environment over a long period may show a carcinogenic effect. Since Mirena is a long-term contraceptive, it would be beneficial for physicians to advise LNG-IUS users about the risk of cancer development. Further studies evaluating the genotoxic effects of Mirena over a long period are needed.

## Data Availability

Data available on request from the authors

## Acknowledgements

This study was carried out with the opportunity provided by the Medical Genetics Department of Meram Medical Faculty.

---

**Figure1.**
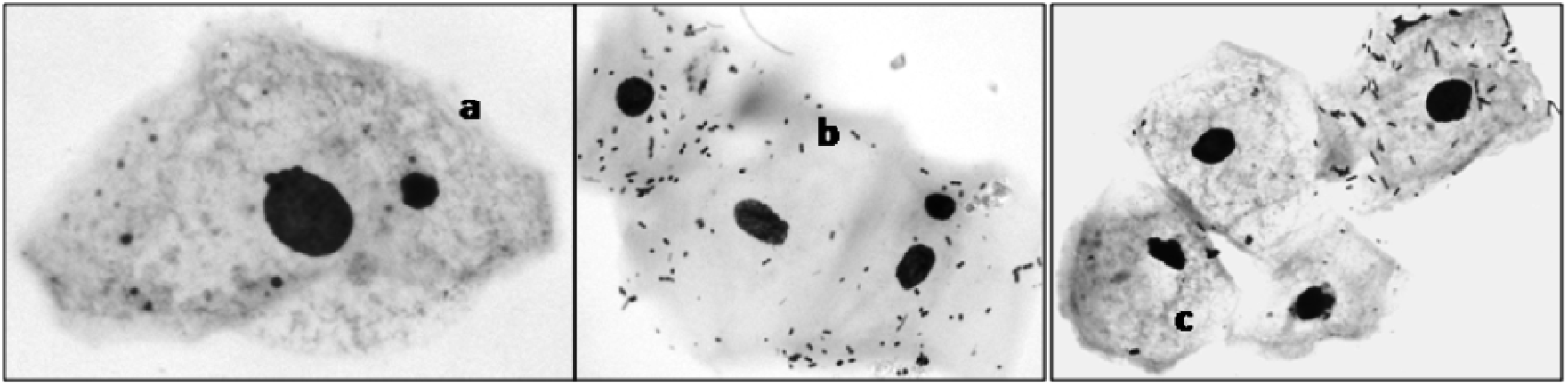
Micronuclei and other nuclear abnormalities observed in cervical cells (a)Micronucleus, (b)karyorrhexis,(c)pyknosis.

